# Smartphone-based videoconference visits are easy to implement, effective, and feasible in Crohn’s Disease Patients: A prospective cohort study

**DOI:** 10.1101/2021.03.11.21253359

**Authors:** Hasan Yilmaz, Ali Erkan Duman

## Abstract

**Background:** Crohn’s Disease patients require life-long follow-up resulting frequent hospital visits. Usefulness of telehealth has been established in remote management of Crohn’s disease but mobile technology’s role is missing. We aimed to determine the feasibility and effectiveness of smartphone-based real-time video visits.

**Methods:** We prospectively studied 139 patients either at a traditional (FTF) or online clinic (OLV) in a university hospital between March 2020-September 2020. We measured patients’ satisfactions, disease activity, visit outcomes, socioeconomic parameters, and travel expenses to assess the effectiveness and acceptance of OLV.

**Results:** Satisfaction scores were significantly higher at OLV compared to FTF (89.58±9.93 vs 70.85±18.51, p<0.001). The Cronbach Alpha reliability coefficient of the VSQ9 scale was 0.878. Median 47km travel distance and 49 minutes travel time per visit saved with OLV. An average of US$12.24 potential travel costs were saved per appointment. Eighty-five per cent of the patients met the needs at online visits and did not require a face-to-face visit.

**Conclusions:** Smartphone-based real-time video visit telehealth model for distant management of Crohn’ disease had high acceptance. The model was easy to implement, effective and saved travel cost and time.

## 1. Introduction

Crohn’s disease is an incurable chronic disease characterised by abdominal pain, diarrhoea, fatigue and extraintestinal manifestations. Patients experience flares and remissions and need treatment across the lifespan. It is estimated that approximately 1 million individuals in the USA and 3.0-3.5 million in Europe have IBD (Inflammatory Bowel Disease), indicating a high disease burden in the Western world.[1]

Despite effective therapies, a significant proportion of patients have suboptimal short and long-term outcomes. The obstacles for effective treatment outcomes include insufficient monitoring of symptoms, difficulty in getting timely access to a gastroenterologist, [2] and lead time, defined as the time interval between Crohn’s disease-specific symptom onset and the establishment of final diagnosis, to effective treatment.[3]

Traditionally, healthcare provider (HCPs) monitor and manage their patients face to face at their office. Travel to the hospital, waiting a long time for the outpatient clinic appointment, difficulty in making an urgent appointment during disease flares, and need for medication prescriptions are the possible barriers to getting quality care. Additionally, the SARS COV-2 pandemic raised concerns among IBD patients, especially those on immunosuppressants or biologic agents, thereby possibly causing treatment delays.[4]

Telehealth refers to remotely delivered healthcare between doctors and patients via telecommunication technologies, either audio or video. Personal computers, mobile applications, web pages and, telemonitoring devices have been used for this purpose. Close monitoring of a patient’s symptoms and adherence to medications can foster more rapid treatment initiation, improved disease outcomes and QoL compared to standard medical care.[5] Previously, a telehealth system that consisted of a provider computer, web-based clinician portal and a support server was found to be practical, acceptable to patients and led to improvements in patient satisfaction.[6] A remote consultation program for underserved areas via a secure online platform reduced the time patients needed to wait to consult gastroenterologists and saved on travel costs.[7]Telehealth for the management of IBD, utilising real-time video visits, also enabled a reduction in travel time and overall visit time.[8]

The aim of the study was to evaluate the feasibility, acceptability, patient satisfaction and economic benefits of smartphone video-based telehealth in the management of Crohn’s disease patients

## 2. Study Design and Method

### 2.1 Overview and Settings

The study was a prospective cohort study conducted at Kocaeli University’s Faculty of Medicine. After the Covid-19 virus breakthrough was declared a pandemic, the Turkish national security institution announced that virtual visits would be reimbursed to prevent viral contamination. Therefore, we started a smartphone-based real-time video visit in patients with Crohn’s disease parallel to traditional visits. Patients voluntarily choose to be included either in face-to-face (FTF) or online visits (OLV). We prospectively collected data to evaluate the effectiveness of online visits.

### 2.2 Inclusion and Exclusion Criteria

The study group included patients with documented Crohn’s Disease, based on clinical, endoscopic, and histologic findings and who had been receiving treatment for more than six months. Participation in the study was offered to all consecutive patients based on a current visit over 18 years old. If the patient does not have a smartphone, we have provided one with an internet connection but patients were excluded from the study if they were unable to use smartphones because of blindness, deafness, or mental disorders.

### 2.3 Description of the Study Groups and TELE-Health System

Two gastroenterologists (HY, AED) examined patients either at a OLV or FTF visit in the outpatient’s clinics. As a telehealth tool, we used the WhatsApp business application, which is freely available and encrypted end to end. All participants received a reminder phone call the day before the appointment. Physicians used an android 10 operating system, octa-core, 2.3 GHz CPU, 32 MPf/2.2 1080p camera smartphone. Internet connection was secured over a virtual local area network and provided by a national academic network at a 60Mps data speed.

The online visit format was the same as that of a traditional FTF visit except for the physical examination. The national online medical information management system “e-nabiz” was used to evaluate laboratory, radiologic and pathology reports. Prescriptions were issued using the electronic prescription system “e-reçete,” and the patients got the medications from the nearest pharmacy with their unique passwords provided at the OLV. (Unlike a phone call visit, real-time video visits with smartphones help the physician for a global assessment of the patient and allow patients to share the results of their examinations instantly.) Face to face clinic patients was seen at hospital outpatient clinics, and standard of care was provided. Participants received a phone call from the administrative personnel the day after the visit, and a questionnaire about satisfaction and patient perceptions/preferences was applied. The patients were informed that questionnaires were anonymised and they cannot be identified.

### 2.4 Outcome Measures

We used a validated, publicly available visit–specific satisfaction instrument (VSQ-9) to determine patient satisfaction. The VSQ-9 Questionnaire had nine items to evaluate physician-patient relations, including the patient perception of the HCP’s, technical skills and personal manner, the amount of time spent on the visits, waiting time for obtaining an appointment, waiting time at the office, accessibility of the office location, and quality of telephone service.[9]

Disease activity was evaluated with the Harvey Bradshaw Index (HBI), where scores below 4 meant quiescent disease, and over 5 indicated active disease.[10]

The distance and travel time between patients’ homes and the hospital was calculated using the Google Maps application, considering the means of transport. We assessed education levels, household, video quality, communication preferences and, visit duration, all of which might influence the use of telehealth technology. Parameters such as current medication, smoking status and, visit outcomes that can affect the course of Crohn’s disease were recorded.

### 2.5 Sample Size and Statistical Analysis

Power analysis was performed using Gpower 3.1 to confirm a sample size of 52 detecting 80% power and alpha with 0.05 for telehealth outcomes between OL and FTF visits. All statistical analyses were performed using IBM SPSS for Windows version 20.0 (SPSS, Chicago, IL, USA). Numeric variables were presented depending on a normal distribution with either mean±standard deviation, or median (IQR). Categorical variables were summarised as counts (percentages). Responses to VSQ9 which consists of a five-level scale was transformed linearly as original study suggested(i.e., poor = 0%; fair = 25%; good = 50%; very good = 75%; and excellent = 100%).[9]Comparisons of numerical variables between groups were carried out using independent samples t-test or the Mann Whitney U test. The association between two categorical variables was examined by the Chi-square test. All statistical analyses were carried out with 5% significance, and a two-sided p-value<0.05 was considered statistically significant. Cronbach alpha, Factor Analysis (FA), and Bartlett’s test statistics were used to determine the construct validity of the VSQ9 scale.

### 2.6 Ethical considerations

This study was performed according to the principles of the Declaration of Helsinki. The protocol was reviewed and approved by the Kocaeli University Ethical Committee of Clinical Research (Project number: GOKAEK-2020/7.12. 2020/123).

## 3. Results

### 3.1 Baseline Demographics

Between March 2020 and September 2020, 180 patients were enrolled in the study (Figure 1). However, 36 (40%) participants from the FTF group and 5 (5%) from the OLV group were lost to follow up after recruitment. The reason for high drop out in the face-to-face group was the fear of coronavirus contamination and unwillingness to complete the study questionnaires.

**Fig. 1.**
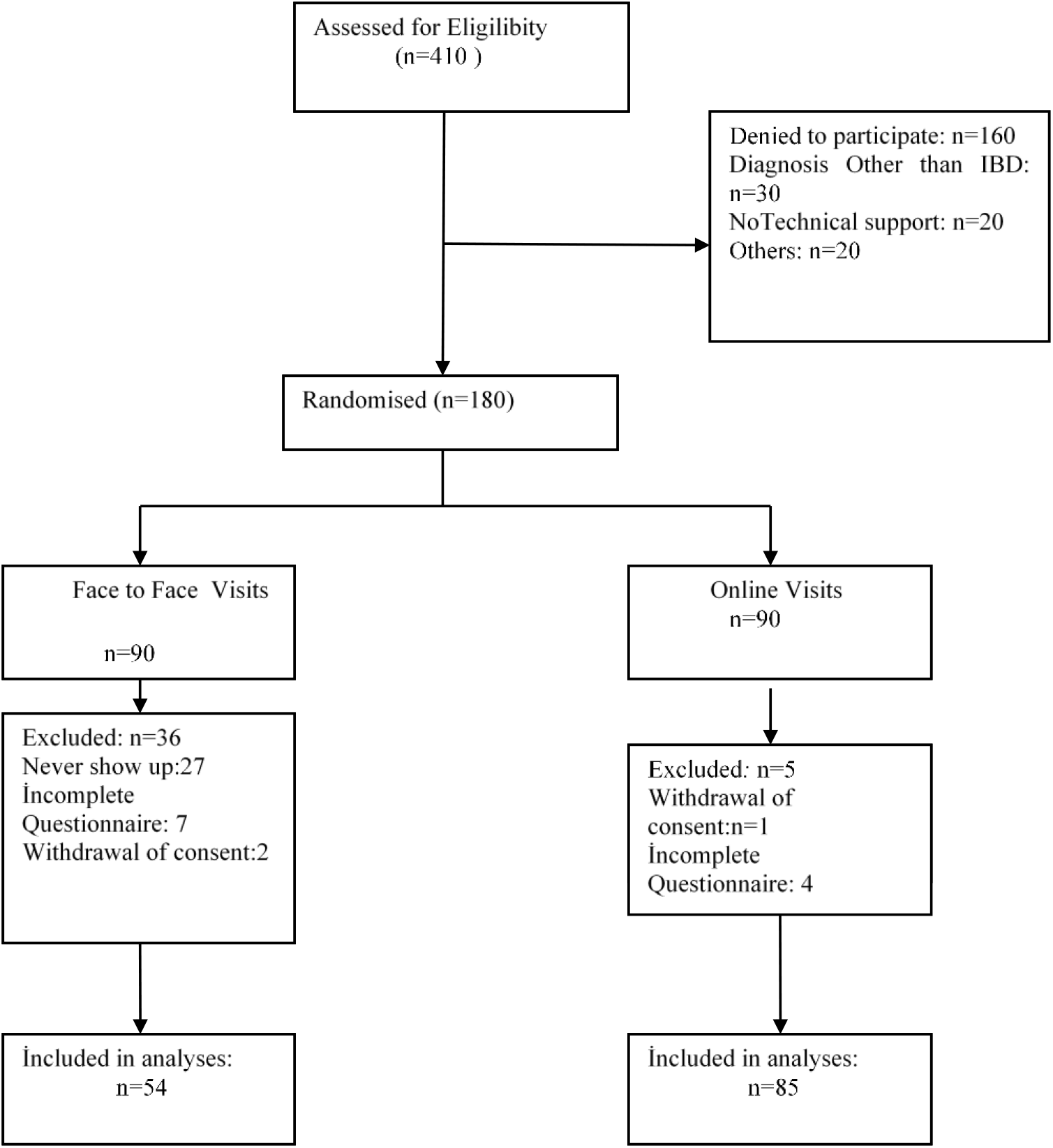
CONSORT Flowchart of the Study

Of the 139 patients included in analyses, the mean age was 45.41 ±13.33 (range 20-79), and 52.5% were women (n=73). There was no significant difference between the clinical and disease characteristics of the OLV and FTF groups. Patients in the FTF group had significantly more Crohn’s Disease complications (p<0.001). Baseline descriptive features of the study population are presented in Table 1.

**Table 1:** Baseline Demographics of the Study Population (n=139)

|  | OLV (n=85) | FTF (n=54) | P Value |
| --- | --- | --- | --- |
| Age<sup>a</sup> | 46.37 ±13.60 | 43.89 ±12.87 | 0.290 |
| Gender,female | 45 (%52.9) | 28(%51.9) | 1.000 |
| Active disease | 27 (%31.8) | 23 (%42.6) | 0.264 |
| **Education Level** |  |  |  |
| low | 41 (%48.2) | 26 (%48.1) | 0.862 |
| high | 44 (%51.8) | 28 % (51.9) |  |
| **Household Income** |  |  |  |
| Low | 36 (%42.3) | 18 (%33.3) | 0.343 |
| Middle | 34 ( %40) | 29 (%53.7) |  |
| High | 15 (%17.7) | 7(% 13) |  |
| **Smoking Status** |  |  |  |
| Non smoker | 36 (%42.4) | 15(% 27.8) | 0.196 |
| Smoker | 29 (%34.1) | 21(% 38.9) |  |
| Quitter | 20 (%23.5) | 18(%33.3) |  |
| **Current Medication** |  |  |  |
| Aminosalicylate | 42 ( %49.4) | 24 ( %44.4) | 0.116 |
| Steroid | 34 (%40) | 28 (%51.8) | 0.164 |
| Azathioprine | 48 (%56.4) | 36 (%66.6) | 0.391 |
| Anti -TNF | 17 (%20) | 18 ( %33.3) | 0.067 |
| Complications | 7 (%8.2) | 19 (%35.2) | 0.001 |
| Disease Duration<sup>a</sup> | 7.40± 5.91 | 5.43 ±4.98 | 0.038 |

Anti-TNF: Anti tumor necrosis factor , <sup>a</sup> Mean (SD)

### 3.2 Satisfaction Scores (VSQ9)

The mean VSQ9 patient satisfaction score of the study population was 86,61±19.25. The mean VSQ9 satisfaction score was significantly higher in the OLV group (89.58 ±9.93) compared to FTF (70.85± 18.51) (p<0.001). Patients in the OLV group were more satisfied in terms of being able to contact the office by phone (OL vs. FTF, 90.77±19.04 vs 47.34±37.37; p<0.001), waiting time for the visit (84,62±19,61 vs. 51,06±25.51;p<0.001), getting an appointment (79,44±20,50 vs 58,51±;p<0.05), time spent with the HCP (91,15±16,78 vs 76,60±24,11; p<0.001), explanation of what was done at the visit (91,92±17,17 vs 80,32±23,27; p<0.05), technical skills (93,85±12,52 vs 79,79±23,68 p<0.05) and the personal manner of the provider (93,08±14,99 vs 86,17±17,13;p<0.05) (Figure 2).The Cronbach Alpha reliability coefficient of the VSQ9 scale was 0.878. Confirmatory Factor Analysis (CFA) was calculated as χ2 = 33,294 (sd = 26; p = 0.154) and RMSEA = 0.051. It was determined that the scale has a high degree of internal consistency and, is valid and reliable.

**Fig. 2.**
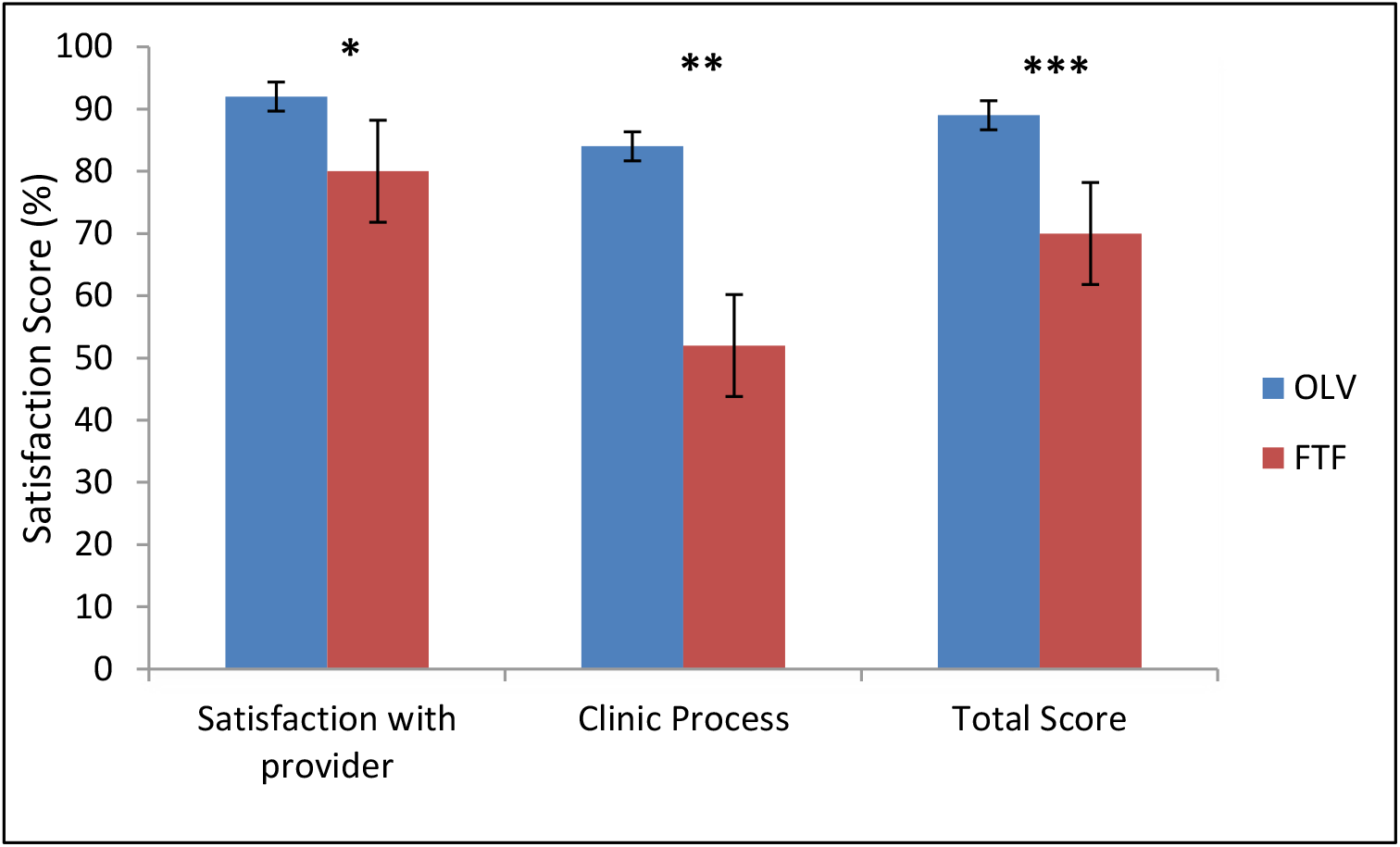
Mean VSQ-9 Satisfaciton Scores OLV: Online visits FTF: Face-to-face visits * p=0.001, ** p< 0.001 *** p< 0.001,

### 3.3 Factors related to the Use of Technology

Participants were asked about their communication activities using communication devices. Text messaging 78 (91%) and video conference calls with friends and relatives 74 (87%) were the most common activities. Thirty-two per cent of the patients reported that they could not undertake video conference visits with their personal computers because of either non-availability of a PC (personal computer) or no confidence to operate. Smartphone ownership rates were not different between OLV patients and FTF patients (82% vs 86% p=0.253)

Patients rated voice and audio quality as good 72 (85.7%), fair 7 (8.3%), and bad 3 (3.6%). Video conference failed with 2 (2.4%) patients because of the poor video and audio quality, where it was impossible to communicate, so we converted to phone visits.

### 3.4 Visit Outcomes

Online visit outcomes were not significantly different from traditional visits. The median visit duration was 12 minutes (Median IQR: 10-14) in the OLV group and 15 (Median IQR: 10-20) minutes in the FTF group. (p<0.05). The outcomes of the visits are summarised in Table 2. Thirteen patients (15.3%) in the OLV group were invited for physical examination due to severe abdominal pain 10 (11.8%), perianal disease 2 (2.4%), and uveitis 1 (1.1%). Sixty-one (71.7%) patients required information about the impact of the Covid19 pandemic on Crohn’s Disease during visits. OLV group patients asked whether they still needed to come to a traditional outpatient clinic after online visits. Seventy-two (85.7%) participants reported no perceived need.

**Table 2:** Outcomes of the Visits.

|  | OLV (n=85) | FTF (n=54) | P-value |
| --- | --- | --- | --- |
| Medication Dose Arrangement | 11(%12.9) | 5 (%11.1) | ns |
| Medication Alteration | 5 (%5.9) | 3 (%5.6) |  |
| Prescription for Refills | 23( %27.1) | 12(%22.2) | ns |
| Smoking Education | 29 ( %31.7) | 20 (%37.1) | ns |
| Laboratory Tests | 10 (24.7) | 14( %25.9) | ns |
| Colonoscopy | 4 (% 4.7) | 3 (%5.5) | ns |
| Hospitalisation | 1 (%1.2) | 2( %3.7) | na |

ns: non spesific na:not applicable

### 3.5 Economic Benefits of the Online Clinic

OLV and FTF participants reported that during the last year, they had visited the hospital for Crohn’s disease a median of four times [ OLV vs FTF, 4(IQR: 2-5 vs 4 (2.25-6); p =0.205]. In our study population, 10 (7.5%) patients had even changed their residence to be closer to a hospital because of frequent visits there. Neither median travel distance to the hospital from patients postal addresses [47 km (IQR: 24-108.5) vs 53 km (IQR (28.5-116.0), p=0.686] nor the travel time [ 49 minutes (IQR: 33.50-98.50) vs, 59 minutes (IQR:36.50 −99.5) p=0.818] was different between OLV and FTF groups. Altogether, the online visits undertaken during the study saved a total travel distance of 10,404 km and 8266-minute travel time compared to traditional visits. An average travel expense per visit of US$12.24 was saved, which constitutes 3% of the 2021 monthly minimum wage (US$385.80) in Turkey.

## 4. Discussion

In the present study, we demonstrated that smartphone-based real-time video visits offered a high-level of satisfaction and acceptance, were easy to implement and decreased travel cost and time.

Given the duration of Crohn’s disease among the study’s participants, their disease activity, percentage of Crohn’s disease complications, socioeconomic spectrum and education levels, our study population can be regarded as quite representative, and satisfaction results can be generalised.

Patient satisfaction with video visits was significantly higher than that of FTF visits. Furthermore, satisfaction scores with the provider and clinical process were found to be greater at online visits. Previously, Krier et al. also reported high patient satisfaction and acceptance of videoconference telemedicine, but they stated that no difference in terms of satisfaction with a PC based telehealth system compared to regular outpatient clinics.[11]Our results differed from the previous trial, possibly because of the use of a more flexible and commonly available technology. We think that OLV were more satisfactory because of the ease of access to a physician who could be consulted on medical problems and possibility of obtaining a prescription by their phone instead of travelling to a hospital. Despite any special clinic time settings, the OLV group had a high satisfaction score with getting an appointment. Mobile technology may give patients more flexibility to join a visit that a hospital offers from anywhere. They may more satisfy getting an appointment easily by this way. In fact, eHealth technologies such as web or text messages are mostly artificial. Online real-time communication with a health care professional could be more realistic and hence increase acceptance.

Users’ cooperation with technology is one of the most prominent hurdles of implementing telehealth. It is critically important to choose the right technological instrument when designing a telehealth model. Ninety-eight per cent of adults in Turkey use a mobile phone, while 77% use smartphones. Among smart telephone users, WhatsApp is currently available on 87.1% of them.[12]Con et al. evaluated IBD patients’ eHealth perspectives and revealed that patients under 30 years old reported higher levels of confidence in using information and communication technologies.

There was an inverse correlation between age and the use of smartphone apps.[13]Additionally, computer anxiety has been found to have a strong negative effect on the acceptance of telehealth services among seniors aged 50 or more.[14]The mean age of our study population was slightly over 40, and the oldest patient was 69 years old. However, we were able to perform online visits successfully with both young and old patients. Entering data with a computer or smartphone app could be challenging, especially for older patients, but real-time video communication to physicians was effortless. The potential loss of privacy with a PC during work time might be another difficulty for the working age population. On the contrary, mobile technology is usable anytime and anywhere.

The online clinic led model of care was also found to bring benefits for socioeconomically disadvantaged patients. Patients with both lower levels of education and low incomes successfully completed the online visits. In contrast to Cross et al.’s recent remote management model for IBD patients, which required a technical support line for participants and providers,[15] participants in our study required neither theoretical and practical education nor technical support to conduct their visits. Real-time videoconferencing using a smartphone was found to be both feasible and easy to implement.

Online clinics saved an average of 47 km travelling distance and a median of 49 minutes travelling time per visit. When we add the waiting time at the office, and the interview itself, patients in the OLV group saved at least half a day. Our results were consistent with the findings of Ruf et al.[8]They also reported that the video conference clinic model reduced the travel distance (mean 310 km) and time per visit (314 minutes) at a rural setting and saved US$36 per visit. However, we do not think telehealth is only efficient for rural populations. Our telehealth model also showed economic and logistic benefits for patients living in crowded metropolitans. Patients reported that they moved their house closer to the hospital because of frequent hospital visits, which indicates that travelling is a burden. Costs related to the caregiver, food, and parking costs, as well as indirect costs like missed work, should also be considered.

According to one meta-analysis, one-third of the telehealth services using real-time video communication increased costs for the service provider.[16]Telehealth technology consisting of a webpage and provider PC requires elements installed in the patients home. It has considerable costs for designing and operating a webpage. However, since we were able to repurposed existing resources, there were no set-up expenses for our video visit model, such as website design, video conference software and hardware, technical equipment, or physical space.

In our study, 15.3% of the patients had to come to FTF for physical examination or an infusion therapy. İn such cases, OLV could not be considered an alternative to traditional visits. The combination of FTF and OLV may be the best practice and can reduce the burden on outpatient clinics.

Our study has some limitations. It was conducted under the extraordinary circumstances of COVID-19. Since the online clinic model can prevent virus dissemination, patients might express greater satisfaction, resulting in selection bias. Once the pandemic is over, patients’ satisfaction perspectives may change. A hybrid visit model which combines online and traditional visits could be a solution for the demands of patients after COVID-19. One of our study’s weaknesses was the absence of follow-up, but its primary aim was to evaluate acceptance, feasibility, and economic benefits. Future studies are needed to determine the long-term effects of OLV on disease activity QoL and medication adherence.

## 5. Conclusion

A smartphone-based video conference telehealth model does not require home installation or additional costs for implementation; it is accessible anywhere and is easy to use. This clinic model yielded widespread acceptance and good satisfaction rates. OLV also brought economic benefits for the patients. However, the long-term effects on disease activity and course need to be determined. We hope that this study will inspire others to implement telehealth to overcome barriers and deliver quality health care for patients with Crohn’s disease.

## Data Availability

Researchers can reach the dataset of this study publicly. (Yılmaz, Hasan (2021), "Smartphone-Based Videoconferance Telehealth for patients with Crohn's Disease", Mendeley Data, V1,

http://dx.doi.org/10.17632/c5d3v8fd5r.1)

## References

1. Burisch J, Jess T, Martinato M, Lakatos PL. The burden of inflammatory bowel disease in Europe. J Crohn’s Colitis [Internet]. 2013 May;7(4):322–37.

2. Borren NZ, Conway G, Tan W, Andrews E, Garber JJ, Yajnik V, et al. Distance to Specialist Care and Disease Outcomes in Inflammatory Bowel Disease. Inflamm Bowel Dis. 2017 Jul;23(7):1234–9.

3. Danese S, Fiorino G, Peyrin-Biroulet L. Early intervention in Crohn’s disease: Towards disease modification trials. Gut [Internet]. 2017 Dec 1 [cited 2020 Dec 11];66(12):2179–87.

4. Mosli M, Alourfi M, Alamoudi A, Hashim A, Saadah O, Al Sulais E, et al. A cross-sectional survey on the psychological impact of the COVID-19 pandemic on inflammatory bowel disease patients in Saudi Arabia. Saudi J Gastroenterol [Internet]. 2020;26(5):263.

5. Elkjaer M, Shuhaibar M, Burisch J, Bailey Y, Scherfig H, Laugesen B, et al. E-health empowers patients with ulcerative colitis: a randomised controlled trial of the web-guided ‘Constant-care’ approach. Gut [Internet]. 2010 Dec 1;59(12):1652LP–1661.

6. Cross RK, Finkelstein J. Feasibility and acceptance of a home telemanagement system in patients with inflammatory bowel disease: a 6-month pilot study. Dig Dis Sci. 2007 Feb;52(2):357–64.

7. Habashi P, Bouchard S, Nguyen GC. Transforming Access to Specialist Care for Inflammatory Bowel Disease: The PACE Telemedicine Program. J Can Assoc Gastroenterol. 2019 Dec;2(4):186–94.

8. Ruf B, Jenkinson P, Armour D, Fraser M, Watson AJ. Videoconference clinics improve efficiency of inflammatory bowel disease care in a remote and rural setting. J Telemed Telecare. 2020 Oct;26(9):545–51.

9. Kennedy DM, Robarts S, Woodhouse L. Patients are satisfied with advanced practice physiotherapists in a role traditionally performed by orthopaedic surgeons. Physiother Can. 2010;62(4):298–305.

10. Harvey RF, Bradshaw JM. A simple index of Crohn’s-Desease activity. Lancet [Internet]. 1980 Mar;315(8167):514.

11. Krier M, Kaltenbach T, McQuaid K, Soetikno R. Potential use of telemedicine to provide outpatient care for inflammatory bowel disease. Am J Gastroenterol. 2011 Dec;106(12):2063–7.

12. Gozde U. Webrazzi Summit 2019 [Internet]. [cited 2021 Jan 22]. p. 634. Available from: https://webrazzi.com/2019/10/24/turkiye-mobil-uygulama-kullanici-sayisi-gemius/

13. Con D, Jackson B, Gray K, De Cruz P. eHealth for inflammatory bowel disease self-management - the patient perspective. Scand J Gastroenterol. 2017 Sep;52(9):973–80.

14. Cimperman M, Makovec Brenčič M, Trkman P. Analyzing older users’ home telehealth services acceptance behavior—applying an Extended UTAUT model. Int J Med Inform [Internet]. 2016;90:22–31.

15. Cross RK, Langenberg P, Regueiro M, Schwartz DA, Tracy JK, Collins JF, et al. A Randomized Controlled Trial of TELEmedicine for Patients with Inflammatory Bowel Disease (TELE-IBD). Am J Gastroenterol. 2019 Mar;114(3):472–82.

16. Wade VA, Karnon J, Elshaug AG, Hiller JE. A systematic review of economic analyses of telehealth services using real time video communication. BMC Health Serv Res [Internet]. 2010;10(1):233.

